# Prognostic significance of compound combined peri-operative biomarkers in gastric cancer

**DOI:** 10.1101/2020.04.21.20066332

**Authors:** Arfon Powell, Alexandra Coxon, David Robinson, Osian James, Adam Christian, Ashley Roberts, Wyn Lewis

## Abstract

**Background:** Survival after gastric cancer surgery is largely attributed to tumor biology, neoadjuvant chemotherapy (NAC), and surgical approach, yet other prognostic factors have been reported, including pre-operative systemic inflammatory response (SIR), and Morbidity Severity Score (MSS). The hypothesis tested was that a SIR, MMS, and pathological composite score, would be associated with disease-free (DFS) and overall survival (OS).

**Methods:** Consecutive 358 patients undergoing potentially curative gastrectomy for adenocarcinoma were studied. Complications were defined as a MSS of Clavien-Dindo classification (CDSC) >1. Serum SIR measurements were performed on the day before surgery, and a composite score (CIMpN) (0-3) was developed based on CRP, morbidity, and pN-stage. Primary outcome measures were DFS and OS.

**Results:** Post-operative complications occurred in 138 (38.5%) patients, (8 (2.2%) deaths), and was associated with higher CRP (28.3% vs. 15.5%, p=0.003), vascular invasion (55.8% vs. 36.8%, p<0.001), and R1 status (26.1% vs. 9.5%, p=0.001). Five-year DFS and OS were 32.9% and 33.3% for patients with post-operative complications compared with 62.5% and 64.0% in controls (p<0.001). Five-year DFS and OS were 31.4% and 37.3% in patients with raised CRPs compared with 58.5% and 59.5% in controls (p=0.005, p=0.001, respectively). Five-year DFS for CIMpN scores of 0, 1, 2, and 3 were 85.9%, 50.0%, 26.2%, and 15.4% (p<0.001) respectively. On multivariable analysis CIMpN score was independently associated with DFS [HR 3.00, 95% Confidence Interval (CI) 1.90-4.73, p<0.001] and OS [1.93 (1.43-2.59), p<0.001].

**Conclusion:** A novel composite score, CIMpN, based on SIR, MSS and pN-stage, offers important prognostic signals.

## Introduction

In global terms, Gastric Cancer (GC) is the third leading cause of cancer related death, accounting for some 740,000 deaths annually^1^. Surgery remains the only potentially curative treatment, but recurrence and metastasis occur in as many as 40% of patients, and survival remains poor even after potentially curative R0 resection. Moreover, such surgery is complex major in nature, inherently high risk, with operative morbidity of 38%^2^ and mortality cited in the most recent UK National Oesophagogastric Cancer Audits ^3^ to be of the order of 2.5%.

Gastric cancer relapse and survival are largely attributed to tumor biology, aggressiveness, and the radicality of the surgery, but other prognostic factors have also been reported, including a pre-operative systemic inflammatory response^4^ and post-operative morbidity^2,5^. In addition to higher recurrence rates, patients who develop post-operative morbidity, experience longer hospital stays and poor quality of life. Therefore, identifying potentially modifiable confounding factors may help improve surgical outcomes measures, reduce recurrence rates and improve long-term survival.

Tumour related inflammation has been termed a prime and seventh hallmark of cancer^6^. Yet the exact aetiology of the systemic inflammatory response (SIR) is unclear, but may be secondary to poor cardiorespiratory fitness or hostile tumour biology. Moreover, it is plausible that the SIR may be implicated in the development of post-operative complications. With the introduction of pre-habilitation programmes^7,8^, attenuating the SIR may represent a promising target in achieving better early outcomes after surgery by means of reducing morbidity and mortality, shortening durations of hospital stay, improving quality of life, thereby boosting long-term survival.

The aim of this study was to determine if markers of the SIR independently predicts post-operative morbidity severity classification and influence prognosis. The hypothesis was that the SIR would be associated with Clavien-Dindo morbidity severity classification, which in turn would be associated with both disease-free and overall survival.

## Methods

### Patients

In order to test the hypotheses proposed in this study, a single cohort was developed and included patients with radiological TNM stage I to III, who following staging were deemed to have potentially resectable gastric adenocarcinoma between January 2009 and August 2018. All patients were managed by a multidisciplinary team with an interest in gastric cancer and included surgeons, oncologists, radiologists, anaesthetists and pathologists.^9^ Preoperative staging involved computed tomography (CT) of the thorax, abdomen and pelvis, including staging laparoscopy when considered appropriate, in order to facilitate individually patient tailored management plans. Selective use of neoadjuvant chemotherapy was adopted following publication of the Medical Research Council Adjuvant Gastric Infusional Chemotherapy (MAGIC) Trial ^10^ in the latter part of the study and was prescribed to 45 patients with minimal comorbidities who were deemed to have relatively advanced disease and would benefit from down-staging of the tumour prior to surgery. Chemotherapy was administered for 3 or 4 cycles preoperatively and postoperatively. Each cycle consisted of epirubicin (50 mg/m^2^) by intravenous bolus, cisplatin (60 mg/m^2^) as a 4-hour infusion on day one and 5-fluorouracil (200 mg/m^2^/day) daily by a continuous intravenous infusion.

The type of surgery for GC was determined by the anatomical location of the tumour; subtotal gastrectomy was performed in patients with antral tumours and total gastrectomy was performed in patients with tumours of the cardia (Siewert type III), body and linitis plastic. A modified extended D2 lymphadenectomy (preserving pancreas and spleen where possible) was performed and the operative approach was open in all cases. In 2010 an enhanced recovery after surgery program was introduced, the details of which have been described previously.^11^

Ethical approval was sought from the regional ethics committee, but a formal application was deemed unnecessary, because the study was considered to be a service evaluation of consecutively recruited patients, in whom consent for treatment had already been provided. In keeping with Cardiff & Vale University Health Board policy, this decision is made by the clinical lead for project and the clinical director.

### Clinicopathological characteristics

Laboratory whole white-cell count, neutrophil count, lymphocyte count, platelet counts, CRP and albumin prior to surgery were recorded. Derivate measurements of the SIR consisted of Neutrophil-Lymphocyte ratio (NLR), Neutrophil-Platelet Score (NPS), Platelet-Lymphocyte Ratio (PLR) and the modified Glasgow Prognostic Score (mGPS) were calculated as described previously.^4^ These derivative measurements were dichotomised into low and high groups by 2.5 for NLR, and 150 for PLR.^4^ The NPS was constructed by grouping patients into three cohorts; zero (0) for patients with both normal neutrophil (≤ 7.5 ×10^9^/L) and platelet counts (≤ 400 ×10^9^/L), one (1) for patients with either a high neutrophil (> 7.5 ×10^9^/L) or platelet count (> 400 ×10^9^/L), and two (2) for patients with both high neutrophil and platelet count. Serum levels of CRP (≤ 10mg/l) and albumin (≥ 35 g/l) were grouped according to previously published thresholds.^12^

Tumours were staged using the seventh edition of the AJCC/UICC-TNM staging system.^13^ Pathological factors were recorded from pathology reports issued at the time of surgery and included tumour differentiation, vascular invasion, margin status and the number of lymph nodes with and without metastasis.

Complications were defined prospectively as any deviation from a pre-determined post-operative course within 30 days following surgery. Complications were diagnosed clinically based on observation, examination and supplementary investigations including but not limited to blood testing (haematology and biochemistry), radiology, and microbiology. Once identified, complications were classified according to the Clavien-Dindo Severity Classification (CDSC).^2,14^ Grade I includes patients with any deviation from normal post-operative course. Grade II complications are treated solely by medicinal therapies. Grade III complications require physical intervention. Grade IV complications are deemed life threatening requiring admission to the critical care unit. Grade V represents post-operative death.

Patients were followed up at regular intervals of 3 months for the first year and 6 months thereafter. In the event that patients developed symptoms suggestive of recurrent disease, investigations were undertaken sooner. Follow-up surveillance was conducted for 5 years or until death whichever was sooner. Overall survival was calculated from time of diagnosis to the date of death. Disease-free survival was measured from the date of surgery until the date of recurrence or date of censoring. No patients were lost to follow-up. Death certification was obtained from the Office for National Statistics via Cancer Network Information System Cymru (CaNISC). Patterns of recurrence were defined as locoregional, distant (metastatic), or both locoregional and distant, when both were diagnosed at the same time. The time of recurrence was taken as the date of the confirmatory investigation.

### Statistical analysis

#### Justification of sample size

Sample size calculations were based on a pre-study literature survey of Cancer Research UK^15^, which has been described previously.^4^ This indicated that the baseline 5-year survival rate of patients diagnosed with stage I gastric cancer was expected to be 80%, compared with 60% in patients with stage II gastric cancer, and a 15% difference in survival would be a realistic expectation. Thus, a minimum of 276 patients were to be studied, providing 80% power to detect such a difference with p<0.05.

#### Methods of data analysis

Grouped data were expressed as median (range) and non-parametric methods were used throughout. Patient demographics were analyzed between the treatment modalities by means of χ2 or non-parametric tests including Mann - Whitney U test. These tests were also employed in the analysis of disease recurrence and time to recurrence for the treatment groups. Disease-free survival for all patients was calculated by measuring the interval from surgery to date of recurrence. Overall survival was measured from the date of diagnosis. Cumulative survival was calculated according to the method of Kaplan and Meier; differences between groups were analyzed with the log rank test. Univariable analyses examining factors influencing survival were examined initially by the life table method of Kaplan and Meier, and those with associations found to be significant (p<0.010) were retained in a Cox proportional hazards model using backward conditional methodology to assess the prognostic value of individual variables. All statistical analysis was performed in SPSS® (IBM® SPSS® Statistics v25.0.0.0, IBM Corporation, Armonk, New York, USA) with extension R.

## Results

### Patients, clinicopathological factors and post-operative morbidity

In total, 358 patients were identified who underwent surgery for GC. The complete baseline characteristics of all clinicopathological variables studied can be found in table 1. The median age for patients undergoing resection was 69 years (IQR 61 - 75) with the majority (42.5%) aged between 65 and 75 years. The majority of patients were male (67.9%), had distal cancers (43.6%), and were lymph node positive (55.1%). Neoadjuvant chemotherapy was prescribed to 81 patients (22.6%), and 96 patients (26.8%) received post-operative adjuvant chemotherapy (table 1). There were 138 (38.5%) patients who developed post-operative complications, which were associated with 8 (2.2%) post-operative deaths within 90 days of surgery and 4 (1.1%) within 30 days of surgery. The frequency of CDSC was grade 1 n=21 (5.9%), grade 2 n=65 (18.2%), grade 3 n=28 (7.8%), grade 4 n=16 (4.5%), and grade 5 n=8 (2.2%). During follow-up, 105 patients (29.3%) developed cancer recurrence and 154 patients (43.0%) died. 286 patients (79.9%) were followed up for 5 years or until death (median 60 (IQR 27-60) months) and no patients were lost to follow-up.

**Table 1:** Univariable and multivariable analysis of clinicopathological factors and complication markers;

|  |  | All morbidity |  |  |  | Major morbidity |  |  |  |
| --- | --- | --- | --- | --- | --- | --- | --- | --- | --- |
|  | Cohort frequency | Univariable |  | Multivariable |  | Univariable |  | Multivariable |  |
|  |  | Hazard ratio<br>(95% CI) | p-value | Hazard ratio<br>(95% CI) | p-value | Hazard ratio<br>(95% CI) | p-value | Hazard ratio<br>(95% CI) | p-value |
| Pre-operative factors |  |  |  |  |  |  |  |  |  |
| Age (Years)<br><65 / 66-75 / >75) | 119 / 152 / 87 | 1.07 (0.81 – 1.42) | 0.631 |  |  | 0.88 (0.59 – 1.30) | 0.505 |  |  |
| Gender<br>(Female / Male) | 115 / 243 | 0.79 (0.50 – 1.22) | 0.278 |  |  | 0.97 (0.52 – 1.82) | 0.924 |  |  |
| Tumour site<br>(Proximal / Body / Distal) | 128 / 74 / 156 | 0.81 (0.64 – 1.03) | 0.092 |  | 0.401 | 1.03 (0.74 – 1.43) | 0.875 |  |  |
| Neoadjuvant therapy<br>(No / Yes) | 277 / 81 | 0.75 (0.44 – 1.26) | 0.274 |  |  | 0.58 (0.26 – 1.29) | 0.181 |  |  |
| ASA grade<br>(2 / 3) | 121 / 237 | 1.43 (0.90 – 2.26) | 0.128 |  |  | 1.46 (0.76 – 2.81) | 0.259 |  |  |
| Anaemia<br>(No / Yes) | 183 / 175 | 1.13 (0.74 – 1.73) | 0.581 |  |  | 1.51 (0.84 – 2.74) | 0.171 |  |  |
| White Cell Count<br>(Low / Normal / High) | 17 / 313 / 28 | 0.81 (0.44 – 1.48) | 0.492 |  |  | 1.53 (0.68 – 3.44) | 0.308 |  |  |
| C-Reactive Protein<br>(Normal / High) | 285 / 73 | 2.16 (1.28 – 3.63) | 0.004 | 1.92 (1.12 – 3.31) | 0.019 | 1.94 (1.01 – 3.74) | 0.047 | 1.90 (0.98 – 3.68) | 0.057 |
| Neutrophil-Lymphocyte ratio<br>(Low / High) | 186 / 172 | 0.82 (0.53 – 1.25) | 0.350 |  |  | 0.63 (0.35 – 1.16) | 0.137 |  |  |
| Platelet-Lymphocyte ratio<br>(Low / High) | 176 / 182 | 1.04 (0.68 – 1.59) | 0.855 |  |  | 1.05 (0.58 – 1.90) | 0.866 |  |  |
| Neutrophil-Platelet Score<br>(Low / Intermediate / High) | 292 / 57 / 9 | 1.01 (0.64 – 1.59) | 0.983 |  |  | 1.34 (0.75 – 2.39) | 0.319 |  |  |
| Pathological factors |  |  |  |  |  |  |  |  |  |
| pT stage<br>(pCR / 1 / 2 / 3 / 4) | 1 / 84 / 36 / 127 / 110 | 1.20 (0.99 – 1.45) | 0.065 |  | 0.630 | 1.14 (0.86 – 1.45) | 0.423 |  |  |
| pN stage<br>(0 / 1 / 2 / 3) | 161 / 72 / 71 / 54 | 1.26 (1.04 – 1.52) | 0.019 |  | 0.977 | 0.93 (0.71 – 1.22) | 0.600 |  |  |
| pTNM stage<br>(I / II / III) | 102 / 120 / 136 | 1.29 (0.99 – 1.68) | 0.063 |  | 0.468 | 1.04 (0.72 – 1.49) | 0.844 |  |  |
| Differentiation<br>(Moderate / Poor) | 185 / 173 | 1.17 (0.76 - 1.79) | 0.472 |  |  | 1.18 (0.66 - 2.13) | 0.575 |  |  |
| Vascular invasion<br>(No / Yes) | 200 / 158 | 2.17 (1.40 - 3.34) | <0.001 | 1.85 (1.18 – 2.91) | 0.008 | 1.90 (1.05 - 3.44) | 0.035 | 1.87 (1.03 – 3.40) | 0.041 |
| R status<br>(0 / 1) | 301 / 57 | 3.35 (1.86 – 6.03) | <0.001 | 2.55 (1.38 – 4.71) | 0.003 | 1.74 (0.85 – 3.57) | 0.131 |  |  |
| Post-operative factors |  |  |  |  |  |  |  |  |  |
| Adjuvant therapy<br>(No / Yes) | 262 / 96 | 0.88 (0.52 – 1.49) | 0.640 |  |  | 0.82 (0.39 – 1.72) | 0.599 |  |  |

### Relationships between post-operative complications, systemic inflammation, and clinicopathological factors

Univariable and multivariable binary logistic regression analysis examining the relationship between both all and major morbidity, systemic inflammation and clinicopathological factors are shown in Table 1. There was a higher proportion of morbidity in patients with a proximal cancer (43.5% vs. 30.9%, p=0.027), both low (8.7% vs. 2.3%, p=0.007) and high total WCC (10.1% vs 6.4%, p=0.007), raised CRP (28.3% vs. 15.5%, p=0.003), vascular invasion (55.8% vs 36.8%, p<0.001) and margin positivity (26.1% vs. 9.5%, p=0.001) (supplementary table 1). The relationships between systemic inflammation, clinicopathological factors and major morbidity (CDSC>2) is shown in supplementary table 2. Major morbidity was associated with higher rates of vascular invasion (57.7% vs 41.8%, p=0.033), raised CRP (30.8% vs. 18.6%, p=0.045), and both a raised (19.2% vs 5.9%, p<0.001) and low WCC (11.5% vs 3.6%, p<0.001) (supplementary >table 2).

**Table 2.** The relationship between clinicopathological factors, disease-free and overall survival in patients undergoing potentially curative resection for gastric cancer.

|  | Univariable |  | Multivariable |  | Univariable |  | Multivariable |  |
| --- | --- | --- | --- | --- | --- | --- | --- | --- |
|  | Disease-free survival |  |  |  | Overall survival |  |  |  |
|  | Hazard ratio (95% CI) | p-value | Hazard ratio (95% CI) | p-value | Hazard ratio (95% CI) | p-value | Hazard ratio (95% CI) | p-value |
| Pre-operative factors |  |  |  |  |  |  |  |  |
| Age (Years)<br><65 / 66-75 / >75) | 0.81 (0.62 – 1.05) | 0.110 |  |  | 0.99 (0.80 – 1.23) | 0.944 |  |  |
| Gender<br>(Female / Male) | 0.65 (0.44 – 0.96) | 0.030 |  | 0.216 | 0.91 (0.65 – 1.27) | 0.912 |  |  |
| Tumour site<br>(Proximal / Body / Distal) | 0.82 (0.66 – 1.02) | 0.080 |  | 0.840 | 0.76 (0.63 – 0.92) | 0.004 | 0.77 (0.64 – 0.93) | 0.006 |
| Neoadjuvant therapy<br>(No / Yes) | 1.43 (0.94 – 2.19) | 0.097 |  | 0.060 | 1.36 (0.95 – 1.95) | 0.089 |  | 0.184 |
| ASA grade<br>(2 / 3) | 0.79 (0.53 – 1.17) | 0.235 |  |  | 0.80 (0.58 – 1.10) | 0.174 |  |  |
| Anaemia<br>(No / Yes) | 1.27 (0.87 – 1.87) | 0.220 |  |  | 1.29 (0.94 – 1.77) | 0.115 |  |  |
| White Cell Count<br>(Low / Normal / High) | 0.93 (0.54 – 1.62) | 0.807 |  |  | 1.24 (0.79 – 1.95) | 0.341 |  |  |
| C-Reactive Protein<br>(Normal / High) | 2.10 (1.37 – 3.21) | 0.001 |  | 0.821 | 2.18 (1.54 – 3.09) | <0.001 |  | 0.852 |
| Neutrophil-Lymphocyte ratio<br>(Low / High) | 1.25 (0.85 – 1.83) | 0.262 |  |  | 1.38 (1.01 – 1.89) | 0.047 |  | 0.380 |
| Platelet-Lymphocyte ratio<br>(Low / High) | 1.26 (0.86 – 1.85) | 0.245 |  |  | 1.28 (0.93 – 1.76) | 0.130 |  |  |
| Neutrophil-Platelet Score<br>(Low / Intermediate / High) | 1.19 (0.82 – 1.74) | 0.362 |  |  | 1.17 (0.86 – 1.60) | 0.326 |  |  |
| Pathological factors |  |  |  |  |  |  |  |  |
| pT stage<br>(pCR/ 1 / 2 / 3 / 4) | 2.23 (1.77 – 2.82) | <0.001 | 1.79 (1.35 – 2.39) | <0.001 | 1.99 (1.66 – 2.38) | <0.001 | 1.67 (1.36 – 2.06) | <0.001 |
| pN stage<br>(0 / 1 / 2 / 3) | 1.87 (1.58 – 2.21) | <0.001 |  | 0.948 | 1.77 (1.55 – 2.03) | <0.001 |  | 0.897 |
| pTNM stage<br>(I / II / III) | 3.33 (2.43 – 4.58) | <0.001 |  | 0.749 | 2.73 (2.14 – 3.47) | <0.001 |  | 0.605 |
| Differentiation<br>(Moderate / Poor) | 2.06 (1.39 - 3.06) | <0.001 |  | 0.096 | 1.78 (1.29 - 2.45) | <0.001 |  | 0.204 |
| Vascular invasion<br>(No / Yes) | 2.82 (1.91 - 4.18) | <0.001 | 1.88 (1.24 – 2.85) | 0.003 | 2.99 (2.16 - 4.14) | <0.001 | 1.78 (1.26 – 2.50) | 0.001 |
| R status<br>(0 / 1) | 3.29 (2.15 – 5.03) | <0.001 |  | 0.088 | 2.29 (2.08 – 4.25) | <0.001 |  | 0.830 |
| Post-operative factors |  |  |  |  |  |  |  |  |
| Adjuvant therapy<br>(No / Yes) | 1.47 (0.96 – 2.25) | 0.079 |  | 0.463 | 1.40 (0.98 – 2.01) | 0.064 |  | 0.819 |
| Post-operative morbidity<br>(No / Yes) | 1.88 (1.28 – 2.76) | 0.001 |  | 0.070 | 2.29 (1.67 – 3.14) | <0.001 | 0.28 (0.12 – 0.63) | 0.002 |
| Clavien-Dindo classification<br>(0 / 1 / 2 / 3 / 4 / 5) | 1.23 (1.06 – 1.42) | 0.005 |  | 0.613 | 1.43 (1.28 – 1.60) | <0.001 | 1.66 (1.29 – 2.13) | <0.001 |
| CIMpN score<br>(0 / 1 / 2) | 3.00 (2.24 – 4.01) | <0.001 | 3.00 (1.90 – 4.73) | <0.001 | 2.29 (1.92 – 2.74) | <0.001 | 1.93 (1.43 – 2.59) | <0.001 |

The relationship between raised CRP and clinicopathological factors can be found in supplementary table 3. Raised CRP was associated with more advanced pT stage (pT4 27.0 vs 45.2%, p=0.002), more advanced pN stage (pN3 12.6 vs 24.7%, p=0.029), more advanced pTNM stage (stage I 33.3% vs. 9.6%, p<0.001), positive margin status (13.0% vs. 27.4%, p=0.003), and post-operative mortality (1.4% vs. 5.5%, p=0.036). CRP measurements were not associated with increasing age (<66 yr. median 3 (IQR 2-7), 66-75 yr. median 4 (IQR 2-9), >75 yr. median 5 (IQR 2-9), p=0.104), gender (both median 4 (IQR 2-9), p=0.610), or higher ASA grade (both groups median 4 (IQR 2-9), p=0.123).

**Table 3.** The relationship between clinicopathological factors and CIMpN score

|  | CIMpN score 0 | CIMpN score 1 | CIMpN score 2 | p-value |
| --- | --- | --- | --- | --- |
| Pre-operative factors |  |  |  |  |
| Age (years) |  |  |  | 0.247 |
| <65 | 25 (26.0) | 55 (38.5) | 39 (32.8) |  |
| 65-75 | 44 (45.8) | 53 (37.1) | 55 (46.2) |  |
| >75 | 27 (28.1) | 35 (24.5) | 25 (21.0) |  |
| Gender |  |  |  | 0.913 |
| Females | 30 (31.3) | 45 (31.5) | 40 (33.6) |  |
| Males | 66 (68.8) | 98 (68.5) | 79 (66.4) |  |
| ASA |  |  |  | 0.374 |
| 2 | 38 (39.6) | 45 (31.5) | 38 (31.9) |  |
| 3 | 58 (60.4) | 98 (68.5) | 81 (68.1) |  |
| Tumour site |  |  |  | 0.024 |
| Proximal | 24 (25.0) | 54 (37.8) | 50 (42.0) |  |
| Body | 29 (30.2) | 28 (19.6) | 17 (14.3) |  |
| Antrum | 43 (44.8) | 61 (42.7) | 52 (43.7) |  |
| Neoadjuvant therapy |  |  |  | 0.321 |
| No | 78 (81.3) | 105 (73.4) | 94 (79.0) |  |
| Yes | 18 (18.8) | 38 (26.6) | 25 (21.0) |  |
| Perioperative factors |  |  |  |  |
| Lymph node yield<br>(median +/- IQR) | 14 (10-20) | 16 (11-24) | 18 (12-26) | 0.004 |
| pT-stage |  |  |  | <0.001 |
| CR | 0 (0.0) | 1 (0.7) | 0 (0.0) |  |
| I | 50 (52.1) | 30 (21.0) | 4 (3.4) |  |
| II | 10 (10.4) | 12 (8.4) | 14 (11.8) |  |
| III | 23 (24.0) | 58 (40.6) | 46 (38.7) |  |
| IV | 13 (13.5) | 42 (29.4) | 55 (46.2) |  |
| pN-stage |  |  |  | <0.001 |
| 0 | 96 (100.0) | 53 (37.1) | 12 (10.1) |  |
| I | 0 (0.0) | 30 (21.0) | 42 (35.3) |  |
| II | 0 (0.0) | 36 (25.2) | 35 (29.4) |  |
| III | 0 (0.0) | 24 (16.8) | 30 (25.2) |  |
| pTNM stage |  |  |  | <0.001 |
| I | 61 (63.5) | 35 (24.5) | 6 (5.0) |  |
| II | 35 (36.5) | 44 (30.8) | 41 (34.5) |  |
| III | 0 (0.0) | 64 (44.8) | 72 (60.5) |  |
| Differentiation |  |  |  | 0.002 |
| Well – moderate | 60 (62.5) | 78 (54.5) | 47 (39.6) |  |
| Poor | 36 (37.5) | 65 (45.5) | 72 (60.5) |  |
| Vascular invasion |  |  |  | <0.001 |
| No | 79 (82.3) | 72 (50.3) | 49 (41.2) |  |
| Yes | 17 (17.7) | 71 (49.7) | 70 (58.8) |  |
| R-status |  |  |  | <0.001 |
| 0 | 95 (99.0) | 127 (88.8) | 79 (66.4) |  |
| 1 | 1 (1.0) | 16 (11.2) | 40 (33.6) |  |
| Post-operative factors |  |  |  |  |
| Operative mortality |  |  |  | 0.004 |
| No | 96 (100.0) | 142 (99.3) | 112 (94.1) |  |
| Yes | 0 (0.0) | 1 (0.7) | 7 (5.9) |  |
| Adjuvant chemotherapy |  |  |  | 0.072 |
| No | 78 (81.3) | 97 (67.8) | 87 (73.1) |  |
| Yes | 18 (18.8) | 46 (32.2) | 32 (26.9) |  |

### Relationship between systemic inflammatory response, post-operative complications, clinicopathological factors, and survival

Univariable and multivariable Cox proportional hazards models of disease-free and overall survival for systemic inflammation, post-operative morbidity, and clinicopathological findings can be found in table 2. Raised CRP was associated with poor disease-free survival (p<0.001) with a median survival of 34 months in the cohort with high CRP, and not reached in the cohort with low CRP. The five-year DFS was 31.4% in the cohort with high CRP, compared with 58.5% in the cohort with low CRP (p=0.001) (supplementary figure 1). Post-operative morbidity was associated with poorer DFS (supplementary figure 2, median survival not reached in either cohort) with five-year DFS of 32.5% in the cohort in whom morbidity was observed, compared with 62.5% in the cohort without morbidity (p=0.001). Given the association between the SIR and post-operative morbidity, the relationship between CRP and DFS was stratified by complication status (figures 1A and 1B). CRP was not associated with DFS in patients developing post-operative morbidity (figure 1B, p=0.388) with a five-year DFS of 46.2% in the cohort with a raised CRP, compared with 52.3% in the cohort with a normal CRP. In patients who did not develop post-operative morbidity, CRP was significantly associated with DFS (figure 1A, p<0.001). The median survival in the cohort with a high CRP was 27 months and not reached in the cohort with a normal CRP. Five-year DFS was 37.9% in the cohort with a high CRP, compared with 76.3% in the cohort with a normal CRP. CRP was associated with DFS in both pN-ve (figure 1C, p=0.023), and pN+ve (figure 1D, p=0.038) patients. In pN-ve patients, the five-year survival was 90% in patients with normal CRPs, and 75% in patients with the high CRPs. In pN+ve patients, five-year DFS was 48% in the cohort with normal CRP, and 28% in the cohort with high CRP. Similar findings were found on OS analysis (supplementary figure 1).

**Figure 1.**
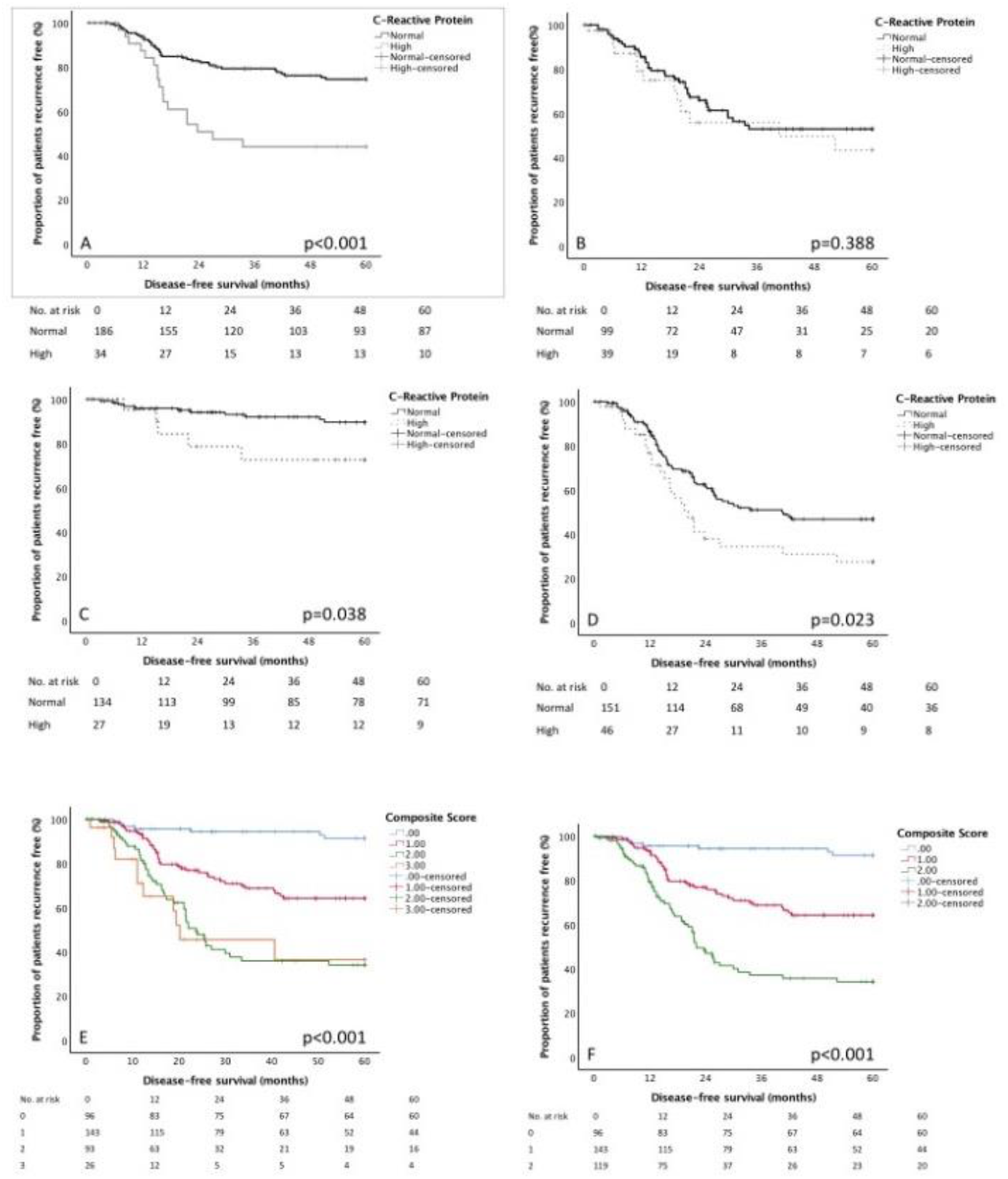
The relationship between pre-operative CRP, disease-free and survival stratified by A.) No postoperative morbidity, B) Post-operative morbidity, C.) pN0 disease, D.) pN1-3 disease, E) CIMpN score (four groups), F) CIMpN score (three groups)

**Figure 2.**
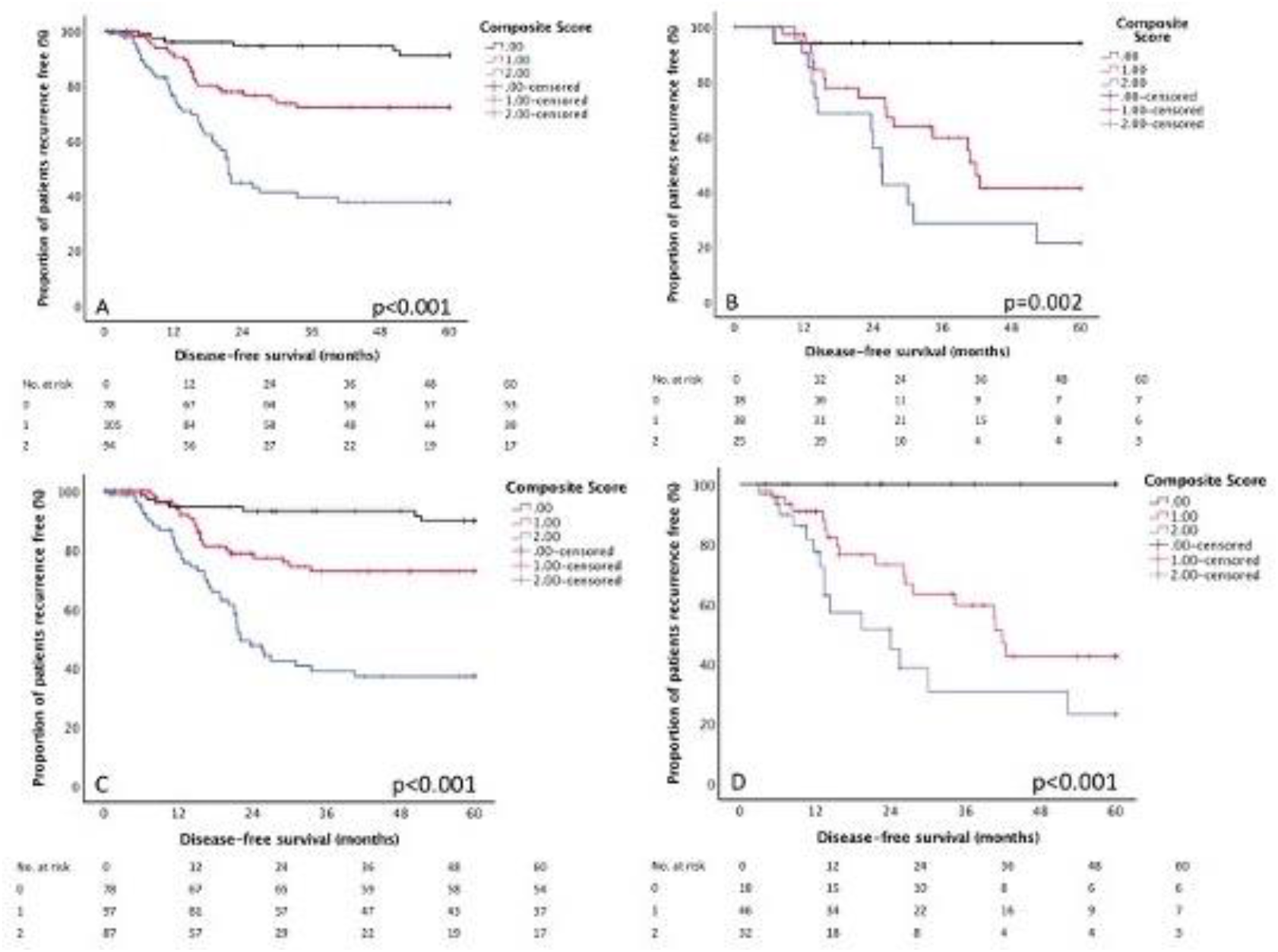
The relationship between disease-free survival and CIMpN score stratified by neoadjuvant and adjuvant chemotherapy status. A.) Did not receive neoadjuvant chemotherapy. B.) Received neoadjuvant chemotherapy. C.) Did not receive adjuvant chemotherapy. D.) Received adjuvant chemotherapy.

Development of a novel composite score encompassing systemic inflammatory response, post-operative morbidity, and pathological lymph node status in gastric cancer

Given the heterogenous survival profiles associated with CRP when stratified by post-operative morbidity and pN-status (figure 1A-D), a novel composite score (CIMpN) was developed based on these variables. Patients were given a cumulative score based on whether they had one of these adverse prognostic factors, thus scores ranged from 0-3. Using this scoring method, 96 (26.8) were classified as a score of 0, 143 (39.9%) a score of 1, 93 (26.0%) a score of 2, and 26 (7.3%) a score of 3. The CIMpN score was associated with poorer DFS (figure 1E, p<0.001); score 0 - DFS was 89.6%, score one - 50.0%, score two - 26.2%, score three - 15.4%. Given the similar survival profiles in patients with scores of two and three these were combined (figure 1F).

### The relationship between CIMpN score, clinicopathological factors, and survival

Univariable and multivariable Cox proportional hazards models for CIMpN score and both DFS and OS can be found in table 2. On multivariable analysis, CIMpN score was associated with poorer DFS (HR 3.00 95%CI (1.90-4.73), p<0.001) and OS (HR 1.93 95%CI (1.43-2.59), p<0.001). The relationship between DFS and novel composite score stratified by neoadjuvant and adjuvant chemotherapy status is shown in figure 2A-D. Similar survival profiles were seen when patients were grouped according to whether or not they had received neoadjuvant treatment (no neoadjuvant, (figure 2A) or adjuvant chemotherapy (figure 2C)) or receiving treatment (received neoadjuvant chemotherapy (figure 2B) or adjuvant chemotherapy (figure 2D)). In patients receiving adjuvant chemotherapy and who had a CIMpN score of zero, five-year DFS was 100%. In patient who did not receive neoadjuvant chemotherapy, CIMpN score (HR 2.34 95%CI (1.63-3.37), p<0.001) was independently associated with DFS on multivariable analysis (supplementary table 4) but not OS (p=0.191).

The relationship between clinicopathological factors and CIMpN score are shown in table 3. Comparing CIMpN scores of zero and two, higher composite score was associated with proximal cancers (25.0% vs. 42.0%, p=0.024), pT4 stage (13.5% vs. 46.2%, p<0.001), poor differentiation (37.5% vs. 60.5%, p=0.002), vascular invasion (17.7% vs. 58.8%, p<0.001), and positive margin status (1.0% vs. 33.6%, p<0.001).

## Discussion

Biomarkers that guide and refine management represent high-value currency in any cancer treatment algorithm. The principal finding of this study was that the SIR, measured using serum CRP, was independently associated with the development of post-operative morbidity and survival. Patients with a raised CRP were nearly twice as likely to develop major post-operative morbidity. Moreover, the differential survival durations observed and associated with critical CRP levels were not mirrored in patients suffering post-operative complications; suggesting that both entities are important independent predictors of survival. The development of a novel composite prognostic score, including Morbidity Severity Score (MSS), and pathological lymph node stage, was associated with both poorer disease-free and overall survival, and moreover was independent of the overall stage of the cancer. In particular, patients scoring zero represented 27% of the cohort and enjoyed disease-free survival of 90%. Administering chemotherapy to patients with scores of one and two may be inappropriate, given the associated poorer survival, and alternative approaches, including pre-habilitation, aimed at optimizing post-operative outcome by reducing or ideally eliminating complications may be more apt.

The SIR has been implicated in the development of post-operative complications following surgery in a number of clinical conditions, including colorectal cancer^16^, heart disease^17,18^, and gastric cancer^19^. Richards *et al*, reported the modified Glasgow Prognostic Score, a composite score of serum CRP and Albumin, was associated post-operative morbidity (Odds Ratio 1.45, p=0.007)^16^. Fransen *et al*, of Maastricht, reported more septic complications (25.3 vs 11.2%, p<0.001) after cardiac surgery, which was also associated with twice as many post-operative deaths (2.7 vs 1.3%)^17^. Perry *et al*, of Boston, USA, reported that CRP dichotomization as low as 3 mg/L was associated with longer durations of hospital stay after coronary artery bypass grafting^18^. Finally, Sato *et al*, of Nagoya, Japan, reported a Systemic Inflammatory Score (SIS), a composite of serum albumin and lymphocyte monocyte ratio, which was strongly associated with a stepwise increase in post-operative morbidity in GC; SIS zero (12%), one (20%), and two (33%, p=0.043)^20^. Ergo, measures to attenuate the SIR prior to surgery, may arguably decrease postoperative morbidity.

The precise trigger of SIR in cancer remains elusive, but may be multifactorial, relating to anatomical degree, tumour biology (aggressiveness), and physiological state, in particular cardiorespiratory fitness. In the present study, patients with a raised CRP had more advanced cancer stage, poorer differentiated tumours, more positive resection margins, and suffered more post-operative mortality. The Physiological and Operative Severity Score for enUmeration of Morbidity and mortality (POSSUM), has been associated with a pre-operative SIR in colorectal cancer^21^. Richards *et al*, from Glasgow, observed that a mGPS of two was observed in 6.4% of patients with the lowest POSSUM score, compared with 25.0% of patients with the highest POSSUM score (p=0.006).^21^ Furthermore, a Malnutrition Universal Screening Tool (MUST) score, based on BMI, weight loss, and active disease was associated with an activated SIR. Almasaudi *et al*, from Glasgow, observed in patients with colorectal cancer, that an mGPS of two was more commonly seen in patients with higher MUST scores, particularly when compared with patients with the lowest scores (8.0% vs 36.0%, p<0.001)^22^. Although confirmatory findings in gastric cancer are unavailable, the available evidence suggests that patients with signals of an activated SIR are vulnerable to more severe post-operative morbidity. Given the underlying components of the CIMpN score are the SIR, morbidity, and lymph node metastasis, it is possible that the best approach of managing this group would be to attenuate the SIR with pre-operative optimization through a prehabilitation programme.

Surgical prehabilitation is a rising and promising arena of research concerned with strategies to optimise patients’ preoperative physical and psychosocial risk profiles, such that postoperative recovery trajectories are boosted, resulting in fewer complications, shorter durations of hospital stay, improved quality of life, and cost-effective prudent health care. Reports so far have focused on a heterogenous group of health interventions, applied within the care continuum, and occurring between diagnosis and the start of surgical treatment. These have included, education, exercise, nutrition, and psychosocial approaches, focused not only the patient, but also the patient’s family, with the aim of promoting health related behavioural change that reaches beyond the immediate preoperative period into the future and longer term. Prehabilitation is the logical precursor to Enhanced Recovery After Surgery Programs (ERP) but should embrace more than just exercise. Nutritional and psychosocial wellbeing are also critical aspects of perioperative care and key components of prehabilitation programs. The preoperative period presents an opportunity to utilize a so-called ‘teachable moment’ and emphasize the importance of positive lifestyle change such as smoking cessation. Future research efforts should explore combining and fusing prehabilitation with ERPs to catalyze additional improvements in outcomes. Moreover, cost-effectiveness evaluation should form part of future research. Prehabilitation in specialties with high risk profiles will probably be associated with additional costs, though it is possible, if not likely, that such costs would be offset by improved outcomes such as shorter durations of hospital stay, fewer complications, and better quality of life. Finally and by tradition, prehabilitation programs are prescriptive and generic; employing a one size fits all philosophy. Bespoke personalized programs, related to individual patients’ physiological, functional, psychosocial profiles, and including combinations of supervised and independent self assessed exercises, delivered in the community rather than secondary care are likely to be associated with greater compliance and effect.

There are a number of inherent limitations and potential criticisms of this study. Primarily, this is a peri-surgical treatment scoring system and a number of barriers exist, that prevent its incorporation into pre-operative treatment planning strategies. Firstly, the perceived radiological lymph node stage is limited, with positive and negative predictive values of 92.9% and 54.7% respectively^23^. Even with additional radiological modalities of CT, EUS and PET CT, nodal size did not predict lymph node metastasis in oesophageal cancer^24^. Pre-operative risk prediction for morbidity severity is also limited, with contradictory results reported related to POSSUM and O-POSSUM^25,26^. Nevertheless, this scoring system has potential in patients receiving primary surgery (uni-modal therapy), who might subsequently be candidates for adjuvant therapy. In contrast, the study has several strengths, benefiting from robust follow-up data with accurate causes and dates of death obtained from the office of national statistics; over 75% were followed up for at least 5 years or death. Patients were recruited consecutively from a single UK geographical region, and had been treated by the same multidisciplinary team and specialist surgeons, using a standardized treatment algorithm, with audited and published quality control. Moreover, surgical outcomes reported in this study compare favorably with national trial and audit data in terms of post-operative outcomes and cumulative survival.^9^

In conclusion, this study demonstrates that the pre-operative Systemic Inflammatory Response, measured with CRP, is associated with post-operative morbidity, and when combined with histopathology lymph node stage, shapes an independent and novel composite score. Although confirmatory evidence is not available in gastric cancer, SIR has been associated with poorer tumour regression grade in response to neoadjuvant chemotherapy in both rectal^27^ and oesophageal cancer^28^. Consequently, SIR may be pivotal in driving cancer progression, post-operative complications, and chemo-resistance. SIR counter measures may offer the best target and multifaceted approach to gastric cancer outcome.

## Data Availability

Queries and data requests can be made to the corresponding author.

## Notes

Conflict of interest – The authors have nothing to declare.

Author Declaration AP - Conceived and designed study, collected data, analyzed data and prepared manuscript. Agreed on final draft for submission. AC - Collected data, helped interpret results, drafted manuscripts and agreed on final draft DR - Collected data, helped with data analysis, drafted manuscript and agreed on final draft. OJ - Collected data, helped with data analysis, drafted manuscript and agreed on final draft. AC - Helped design study and interpret results. Drafted manuscript and agreed on final draft. AR - Helped design study and interpret results. Drafted manuscript and agreed on final draft. WL – Helped conceive and design the study. Helped with data analysis and preparation of manuscript. Agreed on final draft.

### Competing Interest Statement

The authors have declared no competing interest.

### Funding Statement

None

